# Clinical severity of Omicron sub-lineage BA.2 compared to BA.1 in South Africa

**DOI:** 10.1101/2022.02.17.22271030

**Authors:** Nicole Wolter, Waasila Jassat, DATCOV-Gen author group, Anne von Gottberg, Cheryl Cohen

**Affiliations:** Centre for Respiratory Diseases and Meningitis, National Institute for Communicable Diseases (NICD) of the National Health Laboratory Service, Johannesburg, South Africa; School of Pathology, Faculty of Health Sciences, University of the Witwatersrand, Johannesburg, South Africa; Division of Public Health Surveillance and Response, National Institute for Communicable Diseases (NICD) of the National Health Laboratory Service, Johannesburg, South Africa; Right to Care, Centurion, Johannesburg, South Africa; School of Public Health, Faculty of Health Sciences, University of the Witwatersrand, Johannesburg, South Africa

**Keywords:** SARS-CoV-2, Omicron, severity, BA.2, South Africa

## Abstract

Early data indicated that infection with Omicron BA.1 sub-lineage was associated with a lower risk of hospitalisation and severe illness, compared to Delta infection. Recently, the BA.2 sub-lineage has increased in many areas globally. We aimed to assess the severity of BA.2 infections compared to BA.1 in South Africa. We performed data linkages for (i) national COVID-19 case data, (ii) SARS-CoV-2 laboratory test data, and (iii) COVID-19 hospitalisations data, nationally. For cases identified using TaqPath COVID-19 PCR, infections were designated as S-gene target failure (SGTF, proxy for BA.1) or S-gene positive (proxy for BA.2). Disease severity was assessed using multivariable logistic regression models comparing individuals with S-gene positive infection to SGTF-infected individuals diagnosed between 1 December 2021 to 20 January 2022. From week 49 (starting 5 December 2021) through week 4 (ending 29 January 2022), the proportion of S-gene positive infections increased from 3% (931/31,271) to 80% (2,425/3,031). The odds of being admitted to hospital did not differ between individuals with S-gene positive (BA.2 proxy) infection compared to SGTF (BA.1 proxy) infection (adjusted odds ratio (aOR) 0.96, 95% confidence interval (CI) 0.85-1.09). Among hospitalised individuals, after controlling for factors associated with severe disease, the odds of severe disease did not differ for individuals with S-gene positive infection compared to SGTF infection (aOR 0.91, 95%CI 0.68-1.22). These data suggest that while BA.2 may have a competitive advantage over BA.1 in some settings, the clinical profile of illness remains similar.

## MAIN TEXT

The Omicron SARS-CoV-2 variant of concern was first reported in South Africa in mid-November 2021. Early data indicated that infection with Omicron (∼99% BA.1 sub-lineage during this period) was associated with a lower risk of hospitalisation and lower risk of severe illness, once hospitalised, compared to Delta variant infection.^1^ Recently, the BA.2 sub-lineage has increased in many areas globally including South Africa, associated with increases in case numbers in some settings. In South Africa, the BA.2 sub-lineage was first detected on 17 November 2021. From week 49 (starting 5 December 2021), the proportion of BA.2 sub-lineage began to increase, making up 84% (27/32) of all sequenced samples by week 5 (week ending 5 February 2022).^2^ Replacement of BA.1 by BA.2 occurred in a period when SARS-CoV-2 case numbers were declining from the fourth wave peak in South Africa and was associated with a brief increase in case numbers in children of school-going age and slowing of the rate of decline compared to previous waves.

Similar to BA.1, BA.2 is associated with substantial loss in neutralising activity in individuals infected with wild-type SARS-CoV-2 or recipients of mRNA vaccines.^3^ BA.2 has also been associated with increased transmissibility compared to BA.1.^4^ However, data are lacking on the clinical severity of the BA.2 sub-lineage compared to BA.1. We aimed to assess the severity of BA.2 infections compared to BA.1 in South Africa.

Using previously described methods^1^, we performed individual-level data linkage for national data from three sources: (i) national COVID-19 case data, (ii) SARS-CoV-2 laboratory test data for public sector laboratories and one large private sector laboratory, and (iii) DATCOV, which is an active surveillance system for COVID-19 hospital admissions in South Africa (including both incidental and attributable admissions). Case and test data were obtained on 29 January 2022, and DATCOV data on 10 February 2022. The BA.1 sub-lineage contains the 69-70 deletion, which is associated with S-gene target failure (SGTF) when tested using the TaqPath™ COVID⍰19 PCR test (Thermo Fisher Scientific, Waltham, MA, USA). BA.2 lacks this deletion, hence infections with BA.2 are S-gene positive on this assay. In this analysis, restricted to tests performed on the TaqPath™ COVID⍰19 assay, S-gene positive and S-gene target failure (SGTF) infections were considered proxies for Omicron sub-lineages BA.2 and BA.1, respectively.

Two multivariable logistic regression models were generated to assess risk factors for (i) hospitalisation and (ii) severe disease among hospitalised individuals, comparing S-gene positive infections (proxy for BA.2) to SGTF infections (proxy for BA.1). We controlled for factors associated with hospitalisation (age, sex, presence of co-morbidity, province, healthcare sector and prior SARS-CoV-2 infection) and factors associated with severity (age, presence of co-morbidity, sex, province, healthcare sector, number of days between the dates of specimen collection and hospital admission, known prior SARS-CoV-2 infection and SARS-CoV-2 vaccination status) in the respective models. Cases were censored to those with a specimen collected before 20 January 2022, to allow for at least three weeks of follow up. Severity analysis was restricted to admissions that had already accumulated outcomes and all patients still in hospital were excluded. Severe disease was defined as a hospitalised patient meeting at least one of the following criteria: admitted to the intensive care unit (ICU), received any level of oxygen treatment, ventilated, received extracorporeal membrane oxygenation (ECMO), experienced acute respiratory distress syndrome and/or died.

From 1 December 2021 through 29 January 2022, 680,555 SARS-CoV-2 infections were reported. From week 49 (starting 5 December 2021) through week 4 (ending 29 January 2022), the proportion of S-gene positive infections increased from 3% (931/31,271) to 80% (2,425/3,031) (Supplementary figure 1). Among 95,470 samples tested using the TaqPath™ COVID-19 PCR assay, 3.6% of individuals with S-gene positive infection (BA.2 proxy) were hospitalised compared to 3.4% with SGTF infection (BA.1 proxy)(Table 1).

**Table 1.**
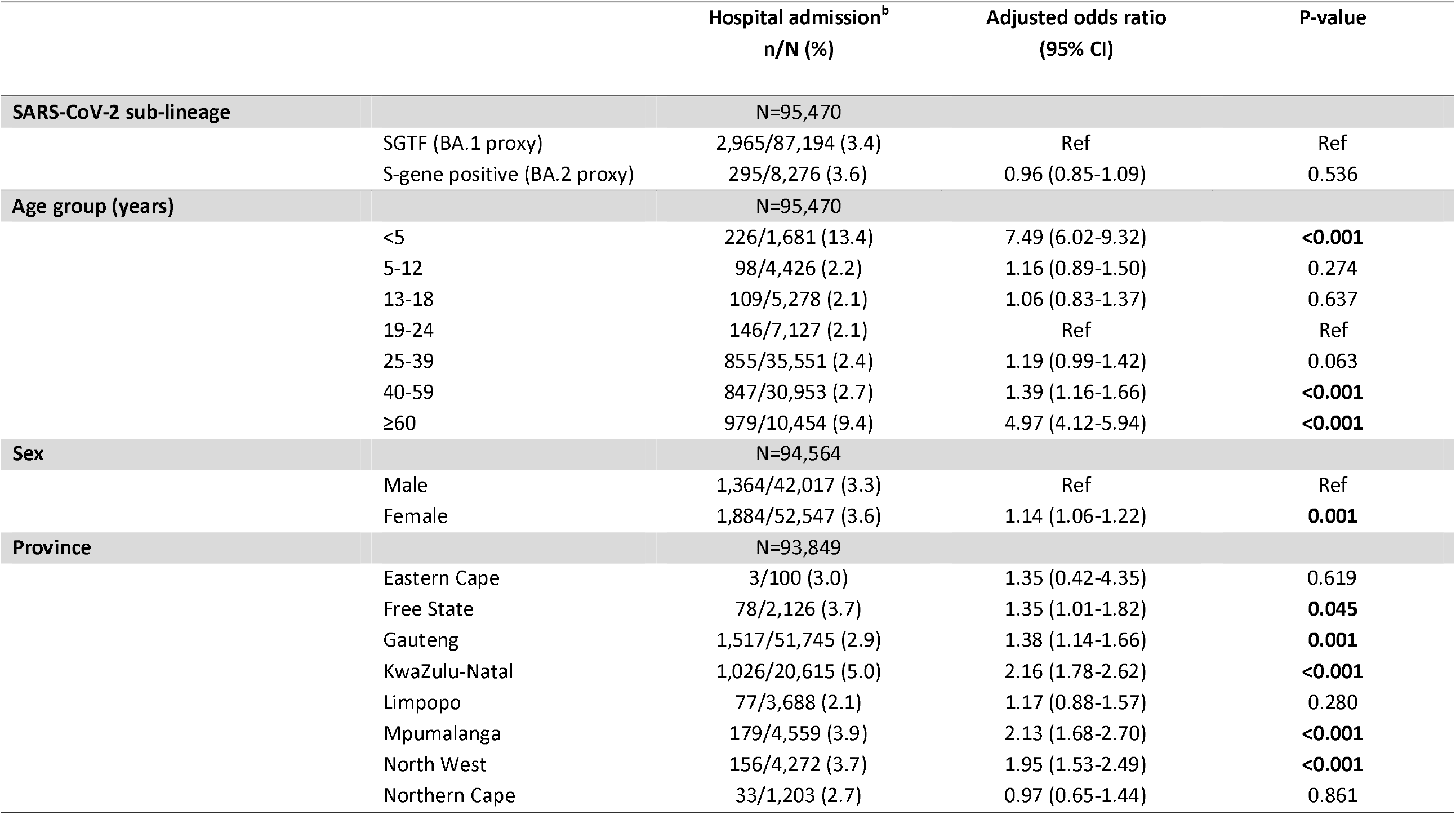
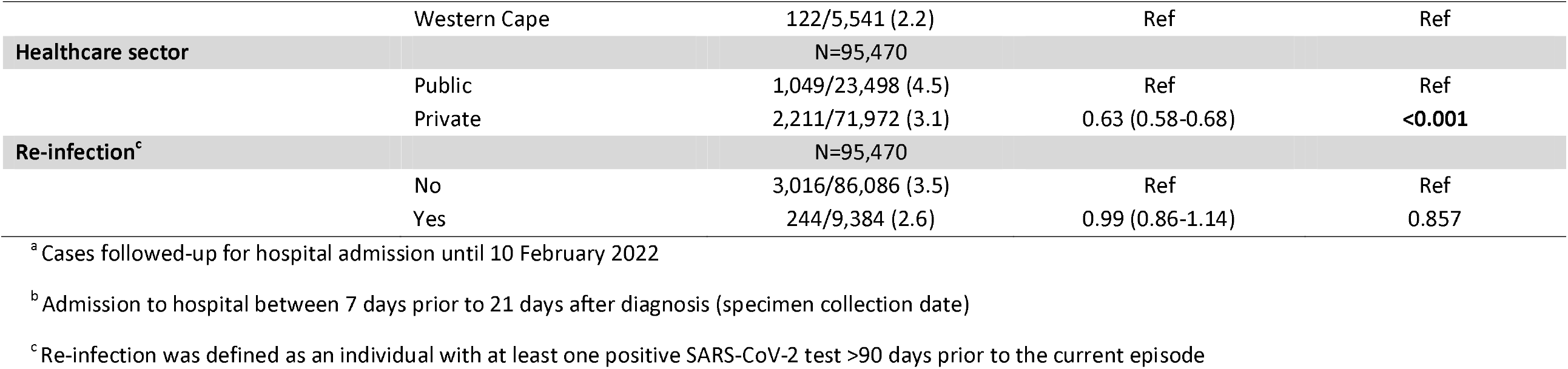
Multivariable logistic regression analysis evaluating the association between S-gene positive infection, compared to S-gene target failure (SGTF) infection, and hospitalisation, South Africa, 1 December 2021 – 20 January 2022^a^ (N=92,962)

|  | Hospital admission<sup>b</sup><br>n/N (%) | Adjusted odds ratio<br>(95% CI) | P-value |
| --- | --- | --- | --- |
| **SARS-CoV-2 sub-lineage** | N=95,470 |  |  |
| SGTF (BA.1 proxy) | 2,965/87,194 (3.4) | Ref | Ref |
| S-gene positive (BA.2 proxy) | 295/8,276 (3.6) | 0.96 (0.85-1.09) | 0.536 |
| **Age group (years)** | N=95,470 |  |  |
| <5 | 226/1,681 (13.4) | 7.49 (6.02-9.32) | <0.001 |
| 5-12 | 98/4,426 (2.2) | 1.16 (0.89-1.50) | 0.274 |
| 13-18 | 109/5,278 (2.1) | 1.06 (0.83-1.37) | 0.637 |
| 19-24 | 146/7,127 (2.1) | Ref | Ref |
| 25-39 | 855/35,551 (2.4) | 1.19 (0.99-1.42) | 0.063 |
| 40-59 | 847/30,953 (2.7) | 1.39 (1.16-1.66) | <0.001 |
| ≥60 | 979/10,454 (9.4) | 4.97 (4.12-5.94) | <0.001 |
| **Sex** | N=94,564 |  |  |
| Male | 1,364/42,017 (3.3) | Ref | Ref |
| Female | 1,884/52,547 (3.6) | 1.14 (1.06-1.22) | 0.001 |
| **Province** | N=93,849 |  |  |
| Eastern Cape | 3/100 (3.0) | 1.35 (0.42-4.35) | 0.619 |
| Free State | 78/2,126 (3.7) | 1.35 (1.01-1.82) | 0.045 |
| Gauteng | 1,517/51,745 (2.9) | 1.38 (1.14-1.66) | 0.001 |
| KwaZulu-Natal | 1,026/20,615 (5.0) | 2.16 (1.78-2.62) | <0.001 |
| Limpopo | 77/3,688 (2.1) | 1.17 (0.88-1.57) | 0.280 |
| Mpumalanga | 179/4,559 (3.9) | 2.13 (1.68-2.70) | <0.001 |
| North West | 156/4,272 (3.7) | 1.95 (1.53-2.49) | <0.001 |
| Northern Cape | 33/1,203 (2.7) | 0.97 (0.65-1.44) | 0.861 |

|  |  |  |  |  |
| --- | --- | --- | --- | --- |
|  | Western Cape | 122/5,541 (2.2) | Ref | Ref |
| **Healthcare sector** |  | N=95,470 |  |  |
|  | Public | 1,049/23,498 (4.5) | Ref | Ref |
|  | Private | 2,211/71,972 (3.1) | 0.63 (0.58-0.68) | **<0.001** |
| **Re-infection<sup>c</sup>** |  | N=95,470 |  |  |
|  | No | 3,016/86,086 (3.5) | Ref | Ref |
|  | Yes | 244/9,384 (2.6) | 0.99 (0.86-1.14) | 0.857 |

<sup>a</sup> Cases followed-up for hospital admission until 10 February 2022
<sup>b</sup> Admission to hospital between 7 days prior to 21 days after diagnosis (specimen collection date)
<sup>c</sup> Re-infection was defined as an individual with at least one positive SARS-CoV-2 test >90 days prior to the current episode

On multivariable analysis, after controlling for factors associated with hospitalisation, the odds of being admitted to hospital did not differ between individuals with S-gene positive (BA.2 proxy) infection compared to SGTF (BA.1 proxy) infection (adjusted odds ratio (aOR) 0.96, 95% confidence interval (CI) 0.85-1.09) (Table 1). In addition to geographic factors, hospital admission was associated with female sex (aOR 1.14, 95% CI 1.06-1.22) and young age (<5 years, aOR 7.49, 95% CI 6.02-9.32) and older age (40-59 years, aOR 1.39, 95%CI 1.16-1.66 and ≥60 years, aOR 4.97, 95% CI 4.12-5.94) compared to individuals aged 19-24 years. Individuals in the private healthcare sector were less likely to be admitted to hospital (aOR 0.63, 95% CI 0.58-0.68) compared to those in the public sector.

Among hospitalised individuals diagnosed from 1 December 2021 to 20 January 2022, after controlling for factors associated with severe disease, the odds of severe disease did not differ for individuals with S-gene positive infection compared to SGTF infection (aOR 0.91, 95%CI 0.68-1.22) (Supplementary table 1). The odds of severe disease was higher among individuals with a comorbidity (aOR 1.52, 95%CI 1.25-1.84) and among individuals aged 40-59 years (aOR 2.09, 95%CI 1.33-3.31) and ≥60 years (aOR 4.36, 95% CI 2.77-6.85), compared to individuals aged 19-24 years. Children aged 5-12 years (compared to 19-24 years), females, and individuals that had received ≥1 SARS-CoV-2 vaccine dose had a lower odds of severe disease.

Limitations of our study include restriction to samples tested with the TaqPath™ COVID-19 PCR assay, biasing data geographically, and that we used S gene positive infection as a proxy for BA.2 sub-lineage infection. Some misclassification could have occurred with other non-Omicron variants, but these made up <2% of all detected viruses in December 2021 and January 2022. There could be a lag in hospitalisation and severe outcomes leading to underestimation of severe illness. To address this we only included hospitalised patients with known outcomes and censored cases to ensure there was at least 3 weeks of follow up. We only had vaccination information for hospitalised cases and this was based on self-report, and re-infection is likely under-ascertained due to limited testing.

We found a similar proportion of individuals were hospitalised and developed severe illness, given hospitalisation, for individuals infected with BA.1 compared to BA.2, during the Omicron-dominated fourth wave in South Africa. These data are reassuring, suggesting that while BA.2 may have a competitive advantage over BA.1 in some settings, the clinical profile of illness remains similar. South Africa may differ from other settings in having a high level of previous immunity following natural infection^5^ and data evaluating BA.2 severity are needed from other settings.

## Data Availability

Data used in this manuscript are available upon reasonable request. Proposals should be directed to.

## Supplementary material

**Supplementary Figure 1.**
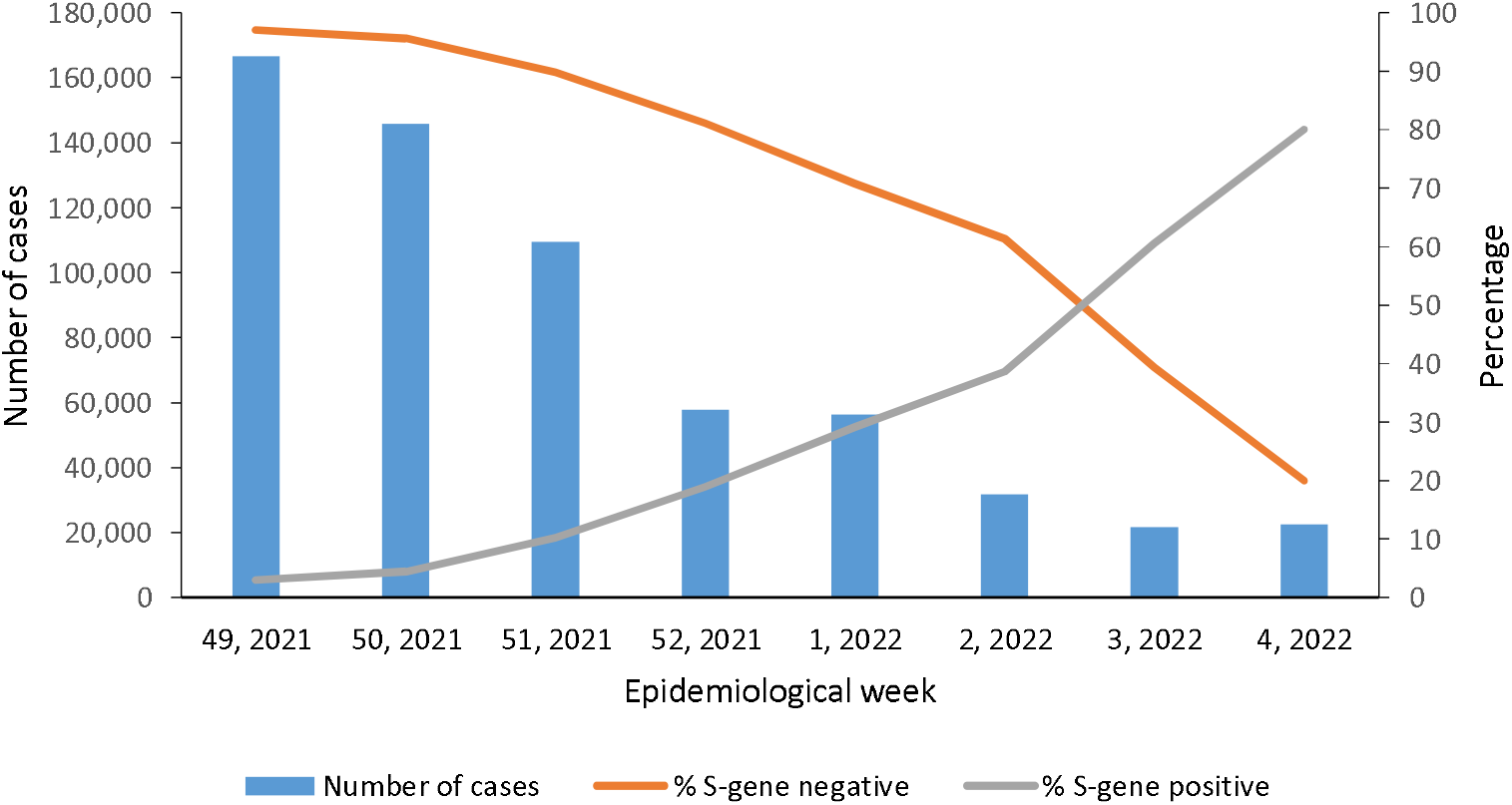
Number of cases detected and percentage of S-gene positive and S-gene target failure (SGTF) infections among tests performed on the TaqPath assay by epidemiological week, DATCOV-Gen, 5 December 2021 – 29 January 2022

**Supplementary table 1.**
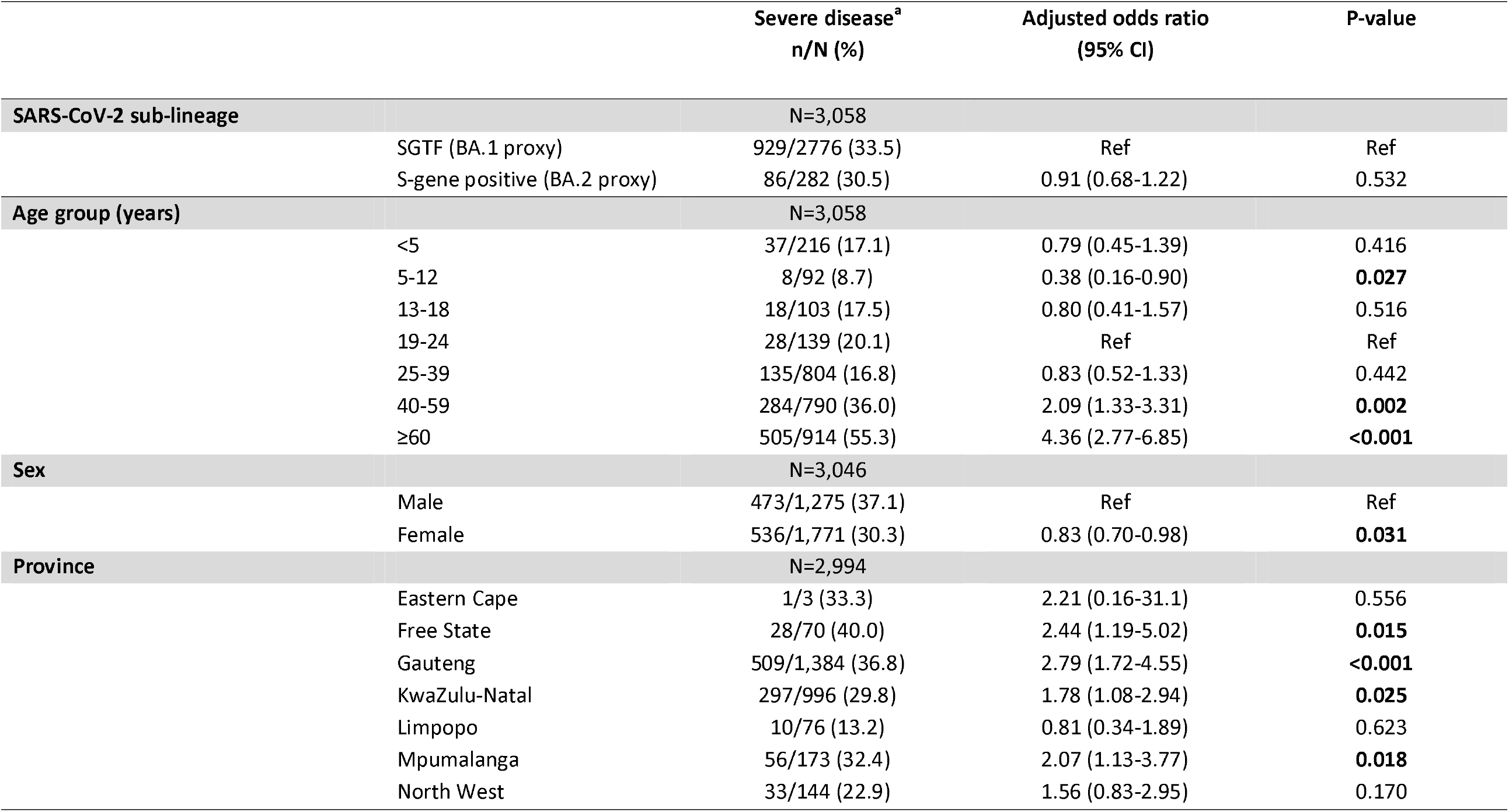
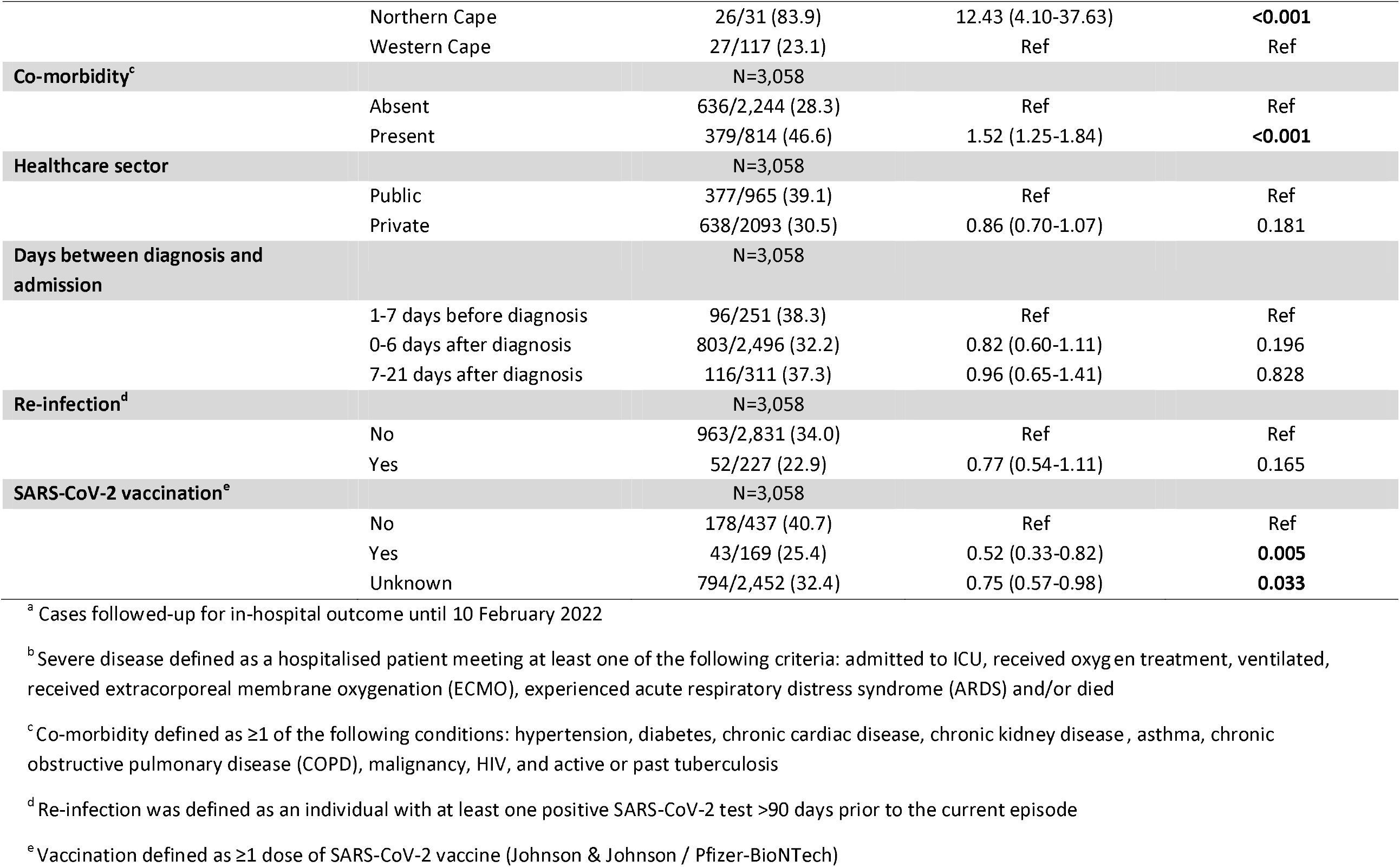
Multivariable logistic regression analysis evaluating the association between S gene positive infection, compared to S-gene target failure (SGTF) infection, and severe disease among hospitalised individuals with known outcome, South Africa, 1 December 2021 – 20 January 2022^a^ (N=2,984)

|  | Severe disease<sup>a</sup><br>n/N (%) | Adjusted odds ratio<br>(95% CI) | P-value |
| --- | --- | --- | --- |
| **SARS-CoV-2 sub-lineage** | N=3,058 |  |  |
| SGTF (BA.1 proxy) | 929/2776 (33.5) | Ref | Ref |
| S-gene positive (BA.2 proxy) | 86/282 (30.5) | 0.91 (0.68-1.22) | 0.532 |
| **Age group (years)** | N=3,058 |  |  |
| <5 | 37/216 (17.1) | 0.79 (0.45-1.39) | 0.416 |
| 5-12 | 8/92 (8.7) | 0.38 (0.16-0.90) | **0.027** |
| 13-18 | 18/103 (17.5) | 0.80 (0.41-1.57) | 0.516 |
| 19-24 | 28/139 (20.1) | Ref | Ref |
| 25-39 | 135/804 (16.8) | 0.83 (0.52-1.33) | 0.442 |
| 40-59 | 284/790 (36.0) | 2.09 (1.33-3.31) | **0.002** |
| ≥60 | 505/914 (55.3) | 4.36 (2.77-6.85) | **<0.001** |
| **Sex** | N=3,046 |  |  |
| Male | 473/1,275 (37.1) | Ref | Ref |
| Female | 536/1,771 (30.3) | 0.83 (0.70-0.98) | **0.031** |
| **Province** | N=2,994 |  |  |
| Eastern Cape | 1/3 (33.3) | 2.21 (0.16-31.1) | 0.556 |
| Free State | 28/70 (40.0) | 2.44 (1.19-5.02) | **0.015** |
| Gauteng | 509/1,384 (36.8) | 2.79 (1.72-4.55) | **<0.001** |
| KwaZulu-Natal | 297/996 (29.8) | 1.78 (1.08-2.94) | **0.025** |
| Limpopo | 10/76 (13.2) | 0.81 (0.34-1.89) | 0.623 |
| Mpumalanga | 56/173 (32.4) | 2.07 (1.13-3.77) | **0.018** |
| North West | 33/144 (22.9) | 1.56 (0.83-2.95) | 0.170 |

|  |  |  |  |  |
| --- | --- | --- | --- | --- |
|  | Northern Cape | 26/31 (83.9) | 12.43 (4.10-37.63) | **<0.001** |
|  | Western Cape | 27/117 (23.1) | Ref | Ref |
| **Co-morbidity<sup>c</sup>** |  | N=3,058 |  |  |
|  | Absent | 636/2,244 (28.3) | Ref | Ref |
|  | Present | 379/814 (46.6) | 1.52 (1.25-1.84) | **<0.001** |
| **Healthcare sector** |  | N=3,058 |  |  |
|  | Public | 377/965 (39.1) | Ref | Ref |
|  | Private | 638/2093 (30.5) | 0.86 (0.70-1.07) | 0.181 |
| **Days between diagnosis and admission** |  | N=3,058 |  |  |
|  | 1-7 days before diagnosis | 96/251 (38.3) | Ref | Ref |
|  | 0-6 days after diagnosis | 803/2,496 (32.2) | 0.82 (0.60-1.11) | 0.196 |
|  | 7-21 days after diagnosis | 116/311 (37.3) | 0.96 (0.65-1.41) | 0.828 |
| **Re-infection<sup>d</sup>** |  | N=3,058 |  |  |
|  | No | 963/2,831 (34.0) | Ref | Ref |
|  | Yes | 52/227 (22.9) | 0.77 (0.54-1.11) | 0.165 |
| **SARS-CoV-2 vaccination<sup>e</sup>** |  | N=3,058 |  |  |
|  | No | 178/437 (40.7) | Ref | Ref |
|  | Yes | 43/169 (25.4) | 0.52 (0.33-0.82) | **0.005** |
|  | Unknown | 794/2,452 (32.4) | 0.75 (0.57-0.98) | **0.033** |

<sup>a</sup> Cases followed-up for in-hospital outcome until 10 February 2022
<sup>b</sup> Severe disease defined as a hospitalised patient meeting at least one of the following criteria: admitted to ICU, received oxygen treatment, ventilated, received extracorporeal membrane oxygenation (ECMO), experienced acute respiratory distress syndrome (ARDS) and/or died
<sup>c</sup> Co-morbidity defined as ≥1 of the following conditions: hypertension, diabetes, chronic cardiac disease, chronic kidney disease, asthma, chronic obstructive pulmonary disease (COPD), malignancy, HIV, and active or past tuberculosis
<sup>d</sup> Re-infection was defined as an individual with at least one positive SARS-CoV-2 test >90 days prior to the current episode
<sup>e</sup> Vaccination defined as ≥1 dose of SARS-CoV-2 vaccine (Johnson & Johnson / Pfizer-BioNTech)

## ETHICAL APPROVAL

Ethical approval was obtained from the Human Research Ethics Committee (Medical) of University of the Witwatersrand for the collection of COVID-19 case and test data as part of essential communicable disease surveillance (M210752), and for the DATCOV surveillance programme (M2010108).

## ACKNOWLEDGEMENTS

We acknowledge all NGS-SA members; laboratory teams at the Centre for Respiratory Diseases and Meningitis and the Sequencing Core Facility of the NICD (Johannesburg, South Africa) for genomic sequencing data; and the national SARS-CoV-2 NICD surveillance team and NICD Information Technology team for NMCSS case data. We thank all laboratories for submitting specimens for sequencing, and all public laboratories and Lancet Laboratories for ThermoFisher TaqPath™ COVID-19 PCR data. We thank all hospitals and health-care workers submitting data through the DATCOV surveillance programme. We are grateful to Cecile Viboud and Kaiyuan Sun of the Fogarty International Center, National Institutes of Health (Bethesda, MD, USA), for their input on the analysis.

## FUNDING STATEMENT

This study was funded by the South African Medical Research Council with funds received from the National Department of Health. Sequencing activities for NICD are supported by a conditional grant from the South African National Department of Health as part of the emergency COVID-19 response; a cooperative agreement between the National Institute for Communicable Diseases of the National Health Laboratory Service and the United States Centers for Disease Control and Prevention (grant number 5 U01IP001048-05-00); the African Society of Laboratory Medicine (ASLM) and Africa Centers for Disease Control and Prevention through a sub-award from the Bill and Melinda Gates Foundation grant number INV-018978; the UK Foreign, Commonwealth and Development Office and Wellcome (Grant no 221003/Z/20/Z); and the UK Department of Health and Social Care and managed by the Fleming Fund and performed under the auspices of the SEQAFRICA project. The Fleming Fund is a £265 million UK aid programme supporting up to 24 low- and middle-income countries (LMICs) generate, share and use data on antimicrobial resistance (AMR) and works in partnership with Mott MacDonald, the Management Agent for the Country and Regional Grants and Fellowship Programme. This research was also supported by The Coronavirus Aid, Relief, and Economic Security Act (CARES ACT) through the Centers for Disease Control and Prevention (CDC) and the COVID International Task Force (ITF) funds through the CDC under the terms of a subcontract with the African Field Epidemiology Network (AFENET) AF-NICD-001/2021. Screening for SGTF at UCT was supported by the Wellcome Centre for Infectious Diseases Research in Africa (CIDRI-Africa), which is supported by core funding from the Wellcome Trust (203135/Z/16/Z and 222754).

The findings and conclusions in this manuscript are those of the author(s) and do not necessarily represent the official position of the funding agencies.

The funders played no role in the writing of the manuscript or the decision to submit for publication.

## DECLARATION OF INTERESTS

CC has received grant support from South African Medical Research Council, UK Foreign, Commonwealth and Development Office and Wellcome Trust, US Centers for Disease Control and Prevention and Sanofi Pasteur. NW has received grant support from Sanofi Pasteur and the Bill and Melinda Gates Foundation. AvG has received grant support from US Centers for Disease Control and Prevention, Africa Centres for Disease Control and Prevention, African Society for Laboratory Medicine (ASLM), South African Medical Research Council, WHO AFRO, The Fleming Fund and Wellcome Trust. RW declares personal shareholding in the following companies: Adcock Ingram Holdings Ltd, Dischem Pharmacies Ltd, Discovery Ltd, Netcare Ltd, Aspen Pharmacare Holdings Ltd. All other authors declare no conflict of interest.

## AUTHOR CONTRIBUTIONS

Conception and design of study: NW, WJ, SW, AvG, CC

Data collection and laboratory processing: NW, WJ, SW, RW, HM, DGA, JE, JNB, CS, NT, NC, MdP, NG, AI, AG, KM, WS, FKT, KS, ZM, NH, RP, JW, AvG, CC

Analysis and interpretation: NW, WJ, SW, RW, HM, MG, DGA, JE, JNB, CS, NC, MdP, NG, AI, AG, KM, WS, FKT, ZM, NH, RP, JW, HH, MD, AB AvG, CC

Accessed and verified the underlying data: NW, RW, HM, DGA, JE, AvG

Drafted the Article: NW, AvG, CC

All authors critically reviewed the Article.

**\*DATCOV-Gen author group:** Sibongile Walaza^1,4^, Richard Welch^3^, Harry Moultrie^2,5^, Michelle Groome^3^, Daniel Gyamfi Amoako^1,6^, Josie Everatt^1^, Jinal N. Bhiman^7,8^, Cathrine Scheepers^7,8^, Naume Tebeila^1^, Nicola Chiwandire^1^, Mignon du Plessis^1,2^, Nevashan Govender^3^, Arshad Ismail^9^, Allison Glass^10^, Koleka Mlisana^11,12^, Wendy Stevens^2,11^, Florette K. Treurnicht^2,11^, Kathleen Subramoney^2, 11^, Zinhle Makatini^2,11^, Nei-yuan Hsiao^11,13^, Raveen Parboosing^2,11,14^, Jeannette Wadula ^2,11,15^, Hannah Hussey^16^, Prof Mary-Ann Davies^16^, Prof Andrew Boulle^16^

^1^ Centre for Respiratory Diseases and Meningitis, National Institute for Communicable Diseases (NICD) of the National Health Laboratory Service, Johannesburg, South Africa

^2^ School of Pathology, Faculty of Health Sciences, University of the Witwatersrand, Johannesburg, South Africa

^3^ Division of Public Health Surveillance and Response, National Institute for Communicable Diseases (NICD) of the National Health Laboratory Service, Johannesburg, South Africa

^4^ School of Public Health, Faculty of Health Sciences, University of the Witwatersrand, Johannesburg, South Africa

^5^ Centre for Tuberculosis, National Institute for Communicable Diseases (NICD) of the National Health Laboratory Service, Johannesburg, South Africa

^6^ School of Health Sciences, College of Health Sciences, University of KwaZulu-Natal, KwaZulu-Natal, South Africa

^7^ Centre for HIV and STIs, National Institute for Communicable Diseases of the National Health Laboratory Service, Johannesburg, South Africa

^8^ SA MRC Antibody Immunity Research Unit, School of Pathology, Faculty of Health Sciences,

University of the Witwatersrand, Johannesburg, South Africa

^9^ Sequencing Core Facility, National Institute for Communicable Diseases of the National Health Laboratory Service, Johannesburg, South Africa

^10^ Lancet Laboratories, Johannesburg, South Africa

^11^ National Health Laboratory Service (NHLS), South Africa

^12^ School of Laboratory Medicine and Medical Sciences, University of KwaZulu Natal, Durban, South Africa

^13^ Division of Medical Virology, University of Cape Town, Cape Town, South Africa

^14^ Department of Virology, University of KwaZulu-Natal, Durban, South Africa

^15^ Department of Clinical Microbiology & Infectious Diseases, CH Baragwanath Academic Hospital, Johannesburg, South Africa

^16^ Western Cape Government: Health and School of Public Health and Family Medicine, University of Cape Town, Cape Town, South Africa

## REFERENCES

1 Wolter N, Jassat W, Walaza S, et al. Early assessment of the clinical severity of the SARS-CoV-2 omicron variant in South Africa: a data linkage study. Lancet 2022; 6736: 1–10.

2 Network for Genomics Surveillance in South Africa (NGS-SA). SARS-CoV-2 Genomic Surveillance Update (11 February 2022). 2022. https://www.nicd.ac.za/wp-content/uploads/2022/02/Update-of-SA-sequencing-data-from-GISAID-11-Feb-2022.pdf (accessed Feb 14, 2022).

3 Iketani S, Liu L, Guo Y, et al. Antibody Evasion Properties of SARS-CoV-2 Omicron Sublineages. bioRxiv 2022; : 2022.02.07.479306.

4 Lyngse FP, Kirkeby CT, Denwood M, et al. Transmission of SARS-CoV-2 Omicron VOC subvariants BA.1 and BA.2: Evidence from Danish Households. medRxiv 2022; : 2022.01.28.22270044.

5 Cohen C, Kleynhans J, von Gottberg A, et al. SARS-CoV-2 incidence, transmission and reinfection in a rural and an urban setting: results of the PHIRST-C cohort study, South Africa, 2020-2021. medRxiv 2021; : 2021.07.20.21260855.

